# Screening for the *alpha* variant of SARS-CoV-2 (B.1.1.7) the impact of this variant on circulating biomarkers in hospitalised patients

**DOI:** 10.1101/2021.06.18.21258699

**Authors:** E. Braybrook, S. Pandey, E. Vryonis, N.R. Anderson, L. Young, D.K. Grammatopoulos

**Author notes:** equal co-authors (E. Braybrook, S. Pandey). Corresponding author: [DKG] Clinical Sciences Research Laboratories, University Hospital of Coventry and Warwickshire; Clifford Bridge Road Coventry CV2 2DX, United Kingdom.

## Abstract

Control of SARS-CoV-2 transmission is complicated by the emergence of variants, especially those containing mutations in the spike protein. By enhancing infectivity and evading immunity, infection with these variants might result in more severe clinical outcomes as well as being more resistant to vaccines developed on the basis of the original prototypic virus variant. One such example is the *alpha* variant (B.1.1.7), which has been detected in more than 100 countries and rapidly become the dominant strain in the UK in late 2020 and early 2021. There is an urgent need to develop appropriate surveillance programmes to rapidly monitor the spread of variants and to better understand the role of variants in disease outcomes and immune evasion. The nucleotide sequencing method, the ‘gold standard’ of variant detection, is unsuitable as a fast-response surveillance tool by frontline diagnostic services which require detection methods with short turnaround times. We developed a screening protocol based of sequential allele-specific qPCR for detection of the N501Y mutation and H69/V70 deletion present in the *alpha*/B.1.1.7 variant. We tested this protocol in previously confirmed positive samples from the Pathology Dept, University Hospital Coventry and Warwickshire during the second wave period in the UK (December 2020-March 2021). In these samples variant identity was confirmed by NGS sequencing via COG-UK. Our results identified increased incidence of variants containing both N501Y and Δ69/70 HV mutations, especially in patients admitted during January and early February 2021. This approach, which yields results within 3 hours, can be used as an initial rapid screening step with NGS as confirmatory follow-up. We also report that the increased prevalence of *alpha*/B.1.1.7 variant in admitted patients since mid-January 2021, a period that characterised peaked mortality rates, was associated with a sharp 2.5-fold rise in the mean circulating IL-6 level and to a lesser extent Troponin-T. More detailed biomarker analysis of a small cohort of patients (n=83), where variant status and clinical outcomes were available, demonstrated that deceased patients infected with the *alpha*/B.1.1.7 variant had significantly higher levels of inflammation and cell injury markers, especially IL-6 and LDH, compared to deceased patients infected with a non-*alpha*/B.1.1.7 variant, pointing towards a more severe inflammatory disease phenotype. In contrast, both groups survivors most biomarker exhibited levels below the group average, with distinct patterns of modified z-scores present.

## INTRODUCTION

As the global pandemic caused by SARS-CoV-2 (1) entered its 2^nd^ year, governments and healthcare systems are still trying to contain and control the spread of the virus. Physical distancing, hygiene, masks, isolation of infected people and their contacts, and aggressive and sometimes prolonged lockdowns, are the main measures employed by governments to minimize different transmission routes and reduce reproductive number (R) and consequently rates of hospital admissions, mortality and morbidities and overall impact on healthcare (2, 3). Since late 2020, vaccines exhibiting encouraging protection rates by preventing SARS-COV-2 infection and transmission, were introduced in various population vaccination programmes. Latest data analysis (4-6) suggest that vaccines have a direct impact by reducing transmission and hospitalization rates, offer real hope that society will be able to re-open and start the long overdue recovery process. However, one aspect of virus biology can potentially hinder these efforts: the emergence of SARS-CoV-2 variants, resulting from continuing viral evolution (7), generated by mutations in key proteins such as the spike protein that alter key characteristics of the virus and potentially exhibit enhanced infectivity/transmissibility and ability to circumvent drug and immune control (8-10). Such mutants might be able to spread more rapidly, cause more severe disease characteristics; decrease effectiveness of the therapeutic agents and evade immunity induced by vaccines or by natural infection. The latter is a major concern because once a large proportion of the population is vaccinated, it is likely immune pressure will develop to favour and accelerate emergence of such variants by selecting for “escape’ mutants.

One such variant is the alpha/B1.1.7 variant (a.k.a. 20I/501Y.V1 Variant of Concern (VOC) 20DEC-01), now confirmed in over 100 countries (11, 12). This variant accrued 23 mutations across the genome including a non-synonymous mutation affecting the receptor binding domain (RBD) of the spike protein at position 501, where the amino acid asparagine (N) has been replaced with tyrosine (Y). Other important mutations, include a H69/V70 deletion, which likely leads to a conformational change in the spike protein which has been associated with viral escape in the immunocompromised; P681H: near the S1/S2 furin cleavage site, a site with high variability in coronaviruses, associated with enhanced membrane fusion of infected cells in *in vitro* experiments.

The emergence of *alpha*/B.1.1.7 variant has been associated with a significant increase in the rate of COVID-19 infections initially in the South East and subsequently in the rest of the United Kingdom, (13, 14). It is suggested that this variant has 40%–80% increased infectivity/transmissibility and early reports raise the possibility of increased risk of death compared with other variants (15-17) and reduced neutralization by some polyclonal and monoclonal antibodies (18). At present it is unclear how this dominant variant can impact effectiveness of public health plans relying upon a rapid protective vaccination program and reduction in transmission through social distancing. Nevertheless, flexible surveillance protocols for detection of the *alpha*/B.1.1.7 are urgently required to monitor prevalence and impact on population heath as well as initiate rapid responses and appropriate containment measures. At present only sequencing the entire virus can be used to detect sequence variations characteristic of the *alpha*/B.1.1.7, however this method, offered only at specialist centres, is not designed to act as a rapid test and alternative approaches are urgently required for implementation in the acute hospital setting. European CDC recommends that multi-target RT-PCR assays that include an S gene target affected by the deletions [resulting in an S gene “drop out” or S gene target failure (SGTF)] can be used as a signal for the presence of the Δ69/70 HV mutation for further investigation (19, 20), especially if sequencing capacity is limited. Many UK Pillar 2 (community testing) labs are using a ThermoFisher TaqPath three-gene nucleic acid amplification test, which is affected SGTF. In fact, since 30 November 2020, 96% of all UK Pillar 2 69-70del sequences were due to the *alpha*/B.1.1.7 lineage (21). However, this approach is not applicable universally as many labs are employing RT-PCR methods that are not affected by deletions of interest.

Recognising this need, we initiated screening of selected positives cases in patients admitted at University Hospital Coventry and Warwickshire (UHCW) NHS Trust during December 2020 to February 2021 by a PCR-based sequential step protocol. We used allele specific RT-PCR to detect simultaneous presence of N501Y and Δ69/70 HV mutations. Positive RNAs were confirmed as *alpha*/B.1.1.7 positive by NGS sequencing. A variation of this approach based on N501Y screening has been used as an early triage step in the Swiss surveillance model (22). We also compared rates of *alpha*/B.1.1.7 positivity with mean levels of routine hospital COVID-19 biomarkers such as IL-6, ferritin, LDH etc and mortality rates across different time intervals to correlate possible impact of *alpha*/B.1.1.7 on disease severity and mortality.

## METHODS

### Study design, patient samples and data and ethics

For the development of the rapid qPCR protocol, deidentified leftover patient nasopharyngeal swab (NPS) samples were used in the study. All patient specimens used were collected in December to February 2021 and previously tested at the Diagnostic Pathology laboratory, UHCW NHS Trust for clinical diagnostic or screening purpose. Specimens were subsequently stored in -80°C freezer until use. A total of 253 positive samples were selected for this study, including samples collected in late December 2020 as well as those collected during January and February 2021. Other than qualitative RT-PCR results (positive or negative), PCR cycle threshold (Ct) values were included in study analysis as well as basic patient metadata including sample date, age, sex, admission to hospital and mortality as well as clinical parameters including laboratory data and patient outcomes. The UHCW NHS Trust COVID-19 research committee exempted this study from ethics oversight as the main purpose was to develop and validate a clinical tool for rapid variant screening and also gather data about emerging variants and laboratory parameters and disease outcomes.

### N501Y+ΔH69/V70 mutation detection by allele specific qPCR

Patient nasopharyngeal swab (NPS) samples were processed as per UHCW NHS Trust routine COVID-19 diagnostic protocols and standard operating procedures. Diagnostic RNA extraction and RT-PCR was carried out using the Abbott RealTi*me* SARS-CoV-2 assay on m2000 platforms. From the RNA extraction final output in 50 μL RNase-free water, 10 μL was used for the variant RT-qPCR reactions. Precautions were taken while handling extracted RNA samples to avoid RNA degradation. Random positive samples were processed for full viral genome sequencing by the University of Birmingham facility of COVID-19 UK Genomics UK Consortium (COG-UK) to identify the viral strains.

The presence of N501Y and ΔH69/V70 mutations was investigated by probe-based melting curve one step RT-PCR assays, using VirSNiP mutation assays by TibMol (#53-0780 and #53-0781, TIB Molbiol, Berlin, Germany) following the manufacturer’s protocol. The testing protocol involved initial assessment of the N501Y mutation followed by assessment of the ΔH69/V70 mutation in N501Y positive samples. Thermal cycling was performed on a Roche LightCycler 2.0 instrument.

Initial validation of the assays established suitability for routine use by determining performance characteristics, analytical sensitivity and specificity, failure rate, minimum RNA quality requirements, repeatability and reproducibility. In each run no-template negative controls were included. In cases of assay failures, testing of samples was repeated.

### Data analysis

Laboratory and clinical data extracted from hospital electronic records were anonymised and analysed to obtain values around number of admissions and hospitalization outcomes over time and average values of specific biomarkers over the same period and two months prior. In total, values from 1,112 blood specimens were analysed. Where multiple results were present for one patient, an overall average was taken. Biomarker values were logged and compared between groups using the independent t-test. Where available, matched *alpha*/B.1.1.7 status and biochemistry parameters were compared to explore any differences associated to *alpha*/B.1.1.7 status. Modified z-scores to examine differences between group scores and the population median were calculated using the following [0.6745*(group value-median of dataset)/median absolute deviation of the dataset]. This was chosen as to evaluate distance of data points from the group median as a more robust comparator than standard z-scores, utilising mean and can be skewed by outliers.

## RESULTS

### RT-PCR performance

Detection of the N501Y mutation was based on melting temperature shifts of the fluorescence peak from 55.9°C (+/- 2.5°C) to 60.55°C (+/- 2.5°C). For the ΔH69/V70 mutation the melting temperature shift was from 56.5°C (+/- 2.5°C) to 63.4°C (+/- 2.5°C) (Fig. 1a). Initial validation demonstrated that both assays exhibited robust reproducibility and repeatability characteristics suitable for routine use (*data not shown*). We tested a wide range of primary RNA samples with Ct values ranging from 15 to 28 (based on the Abbott RealT*ime* SARS-CoV-2 assay) and both assays were able to amplify the target sequence (Fig.1b). In addition, sensitivity and specificity experiments identified that successful amplification was achieved even when the primary sample was diluted by up to 1:100 (*data not shown*), whereas specificity was confirmed by complete concordance between the RT-PCR and NGS results in correctly identifying presence or absence of each mutation. In total 30 samples were processed for both NGS sequencing and RT-PCR.

**Fig.1.**
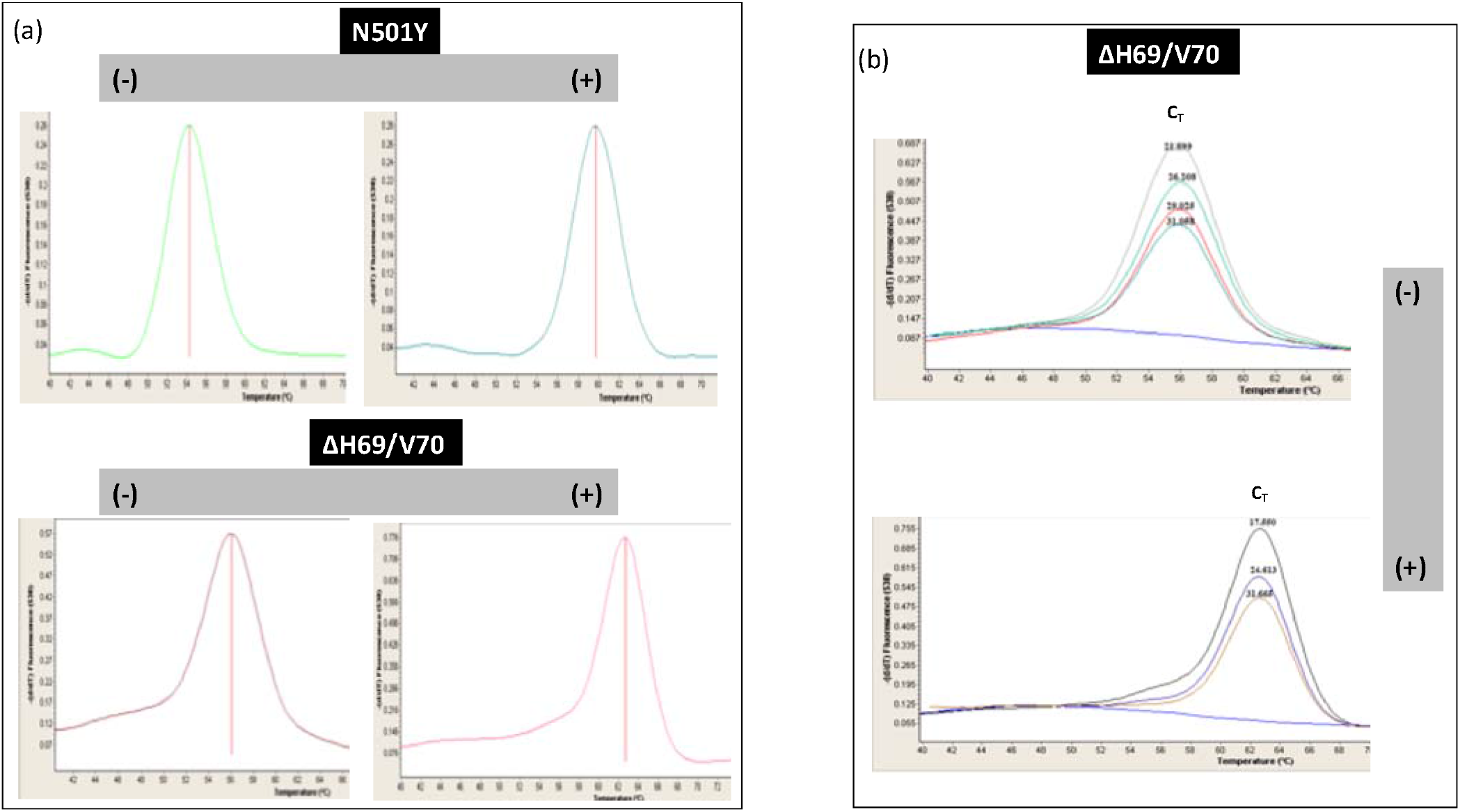
Melting curve analysis of N501Y and Δ69/70 HV mutations present in the B.1.1.7 VOC. SARS-COV2 positive RNAs were investigated by probe-based melting curve one step RT-PCR assays, using VirSNiP mutation assays. (a) Representative melting curve analysis for each mutation investigated are shown ; (b) Additional experiments investigated the impact of viral load on signal intensity for the Δ69/70 HV mutation. *Top*: fluorescence peaks of Δ69/70 HV negative samples with Ct values ranging between 21.7-31.1. *Bottom*: fluorescence peaks of Δ69/70 HV positive samples with Ct values ranging between 17.6-31.7.

As mentioned, 253 RNA samples were processed for mutation screening and successful implementation of the PCR protocol was achieved in 235 samples. In processing real world specimens, we found that the N501Y VirSNiP mutation assay had a failure rate of around 7%, whereas the failure rate of the ΔH69/V70 assay was 2%. No specific issues around primary sample purity or concentration were identified after NanoDrop Nucleic Acid Quantification at wavelengths of interest 260nm, 280nm and 230nm. This suggests that specimen integrity and potential degradation due to incorrect handling and/or storage appears to be an important factor, mostly affecting, the N501Y VirSNiP assay. We also noted that all test failures were on samples with an original PCR detection assay Ct>32 indicating that low viral loads might affect assay performance. In fact, in approximately 30% of samples with Ct>32 PCR failed to amplify the target sequence.

### *Alpha*/B.1.1.7 variant screening during the winter 2020-21 COVID-19 wave

The *alpha*/B.1.1.7 screening protocol was based on sequential detection of N501Y and ΔH69/V70 mutations. Samples positives for the N501Y mutation were then processed for the ΔH69/V70. Most importantly, the short reaction time of each RT-PCR amplification (<1h) allows results to be generated the same day. Interestingly, all RNA samples tested positive for the N501Y were also positive for the ΔH69/V70 mutation (Fig.2), suggesting that, during the period tested (Dec’2020-Feb’2021), the *alpha*/B.1.1.7 was the dominant if not the exclusive strain carrying the N501Y mutation in the region of Coventry and Warwickshire. Although only a small proportion of RNAs (2-7.6%) were available for trialling this protocol, a clear pattern of temporal transmission characteristics emerged, as progressively the majority of samples tested, were positive for the *alpha*/B.1.1.7 VOC. Only 1 out of 40 samples tested in December 2020 was positive for the *alpha*/B.1.1.7 VOC; however, the positivity went up to 60.3% and 93.4% of samples tested during January to February 2021 indicating that during this period when the UK experienced the second COVID-19 wave, the *alpha*/B.1.1.7 VOC became the dominant SARS-CoV-2 variant transmitted around Coventry and Warwickshire. Sequencing data confirmed that samples without N501Y and ΔH69/V70 mutations were of the B1.177 “Spanish’ variant, the major European lineage, characterized by three single nucleotide polymorphisms (SNPs) (C22227T, C28932T, and G29645T) (23).

**Fig.2.**
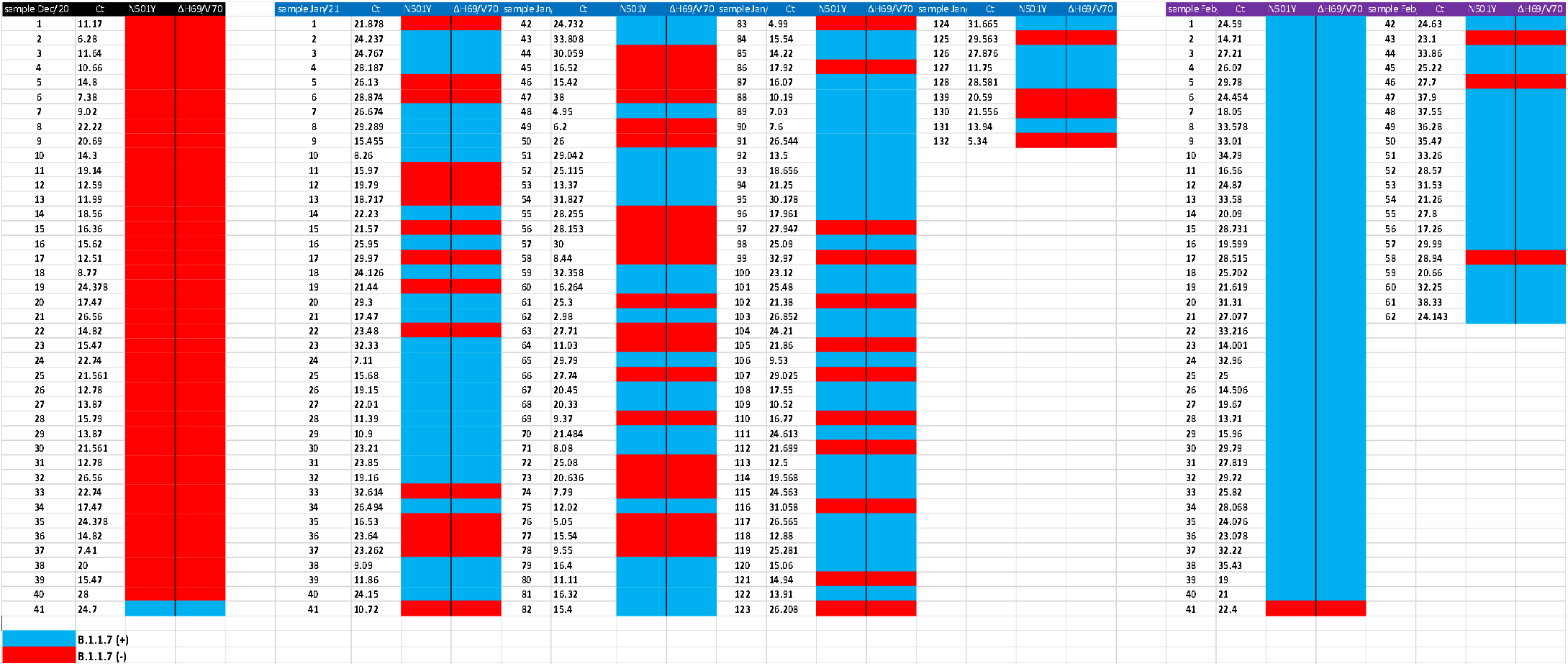
Overview table of SARS-COV2 positive RNA samples processed for detection of N501Y and Δ69/70 HV mutations. *Red* colour depicts samples without any N501Y and Δ69/70HV mutations, whereas blue colour shows samples positive for N501Y and Δ69/70HV mutations. The Ct values obtained from the diagnostic RT-PCR are also shown.

### Temporal patterns of COVID-19 biomarker levels

The increased prevalence of variants containing mutations affecting the RBD that potentially alter pathogenic characteristics and severity of viral infection prompted us to examine changes in mean levels of blood biomarkers of laboratory-confirmed COVID-19 patients, measured in the Biochemistry laboratory during the second wave (Dec’2020-March2021) or the period that immediately preceded this (Oct-Nov’2020). Since the start of the pandemic these biomarkers have been used in the routine care of symptomatic COVID-19 in-patients (24), mainly as a tool of assessing disease severity. Analysis of mortality rates in relation to admissions during the period Oct’2020-Feb2021 identified an increase in mortality rate from 6% in early October to over 20% during the first week of December 2020 that peaked during the second half of January 2021 (rate >27%) and then progressively remained between 15-19% until the first week of March 2021 (*data not shown*).

Plotting the averages of each biomarker over time (Table 1), expressed as the percentage change to values obtained at time 0 (start of Oct 2020) demonstrated comparable pattern of temporal variation for most biomarkers associated with small transient changes (Fig.3). Interestingly three biomarkers, IL-6, BNP and cTnT exhibited distinct patterns, characterised by a transient increase during the first half of December 2020 and subsequently a sustained rise in levels during the second half of January 2021 that remained high during February 2021. Levels of IL-6 n particular were disproportionately high during the period (Jan-Feb2021) reaching maximal values that were 2-2.5 x above baseline. In fact, our analysis demonstrated that although the average of IL-6 levels measured during a 14-week period up to 10^th^ of January 2021 was 128± 489 ng/L, this was significantly increased by 71% during the next 8-weeks up to 7/3/21. Average levels of markers of cardiac function cTroponin T (cTnT) and NT-pro-BNP showed similar trends albeit not significant, whereas other inflammatory markers such as CRP, ferritin and LDH did not follow this pattern.

**Table 1:** Mean±SD values of specific blood biomarkers measured in COVID-19 inpatients during the period 5/10/20-10/1/2021 and 11/1/2021-7/3/2021. Symbols and units in brackets: PCT: procalcitonin (ug/l); IL-6: interleukin-6 (ng/l); CRP: C-reactive protein (mg/l); FER: ferritin (ug/l); TRN: transferrin (g/l); Iron (umol/l); ALBU: albumin (g/l); LDH: lactate dehydrogenase (U/l); TNT: troponin T (ng/l); NT-PROBNP: N-terminal pro B-type natriuretic peptide (pmol/l); HB: haemoglobin (g/l); N/L: neutrophil to lymphocyte ratio; SII: systemic immune-inflammation index * *p*<0.05 between groups

| Laboratory test -dates |  | PCT | IL-6* | CRP | FER | TRN | IRON | ALBU | LDH | TNT | NT-PRO BNP | HB | N/L | SII |
| --- | --- | --- | --- | --- | --- | --- | --- | --- | --- | --- | --- | --- | --- | --- |
| Oct 5th - Jan 10th | Mean | 1.95 | 128 | 88 | 1084 | 1.8 | 8.6 | 33.3 | 673.5 | 37.7 | 277.8 | 122.6 | 10.6 | 2854.2 |
|  | SD | 8.79 | 490 | 88 | 1238 | 0.5 | 6.4 | 6 | 358.4 | 82.3 | 650.1 | 22.6 | 9.4 | 2877.2 |
| Jan 11th - March 7th | Mean | 1.14 | 220 | 77 | 1193 | 1.9 | 9.7 | 34.1 | 713.8 | 50.4 | 299.6 | 121.1 | 11 | 2988.3 |
|  | SD | 4.59 | 588 | 82 | 2610 | 0.6 | 5.8 | 6.8 | 458.8 | 153.4 | 724.8 | 23.3 | 11.1 | 3270.1 |

**Fig.3.**
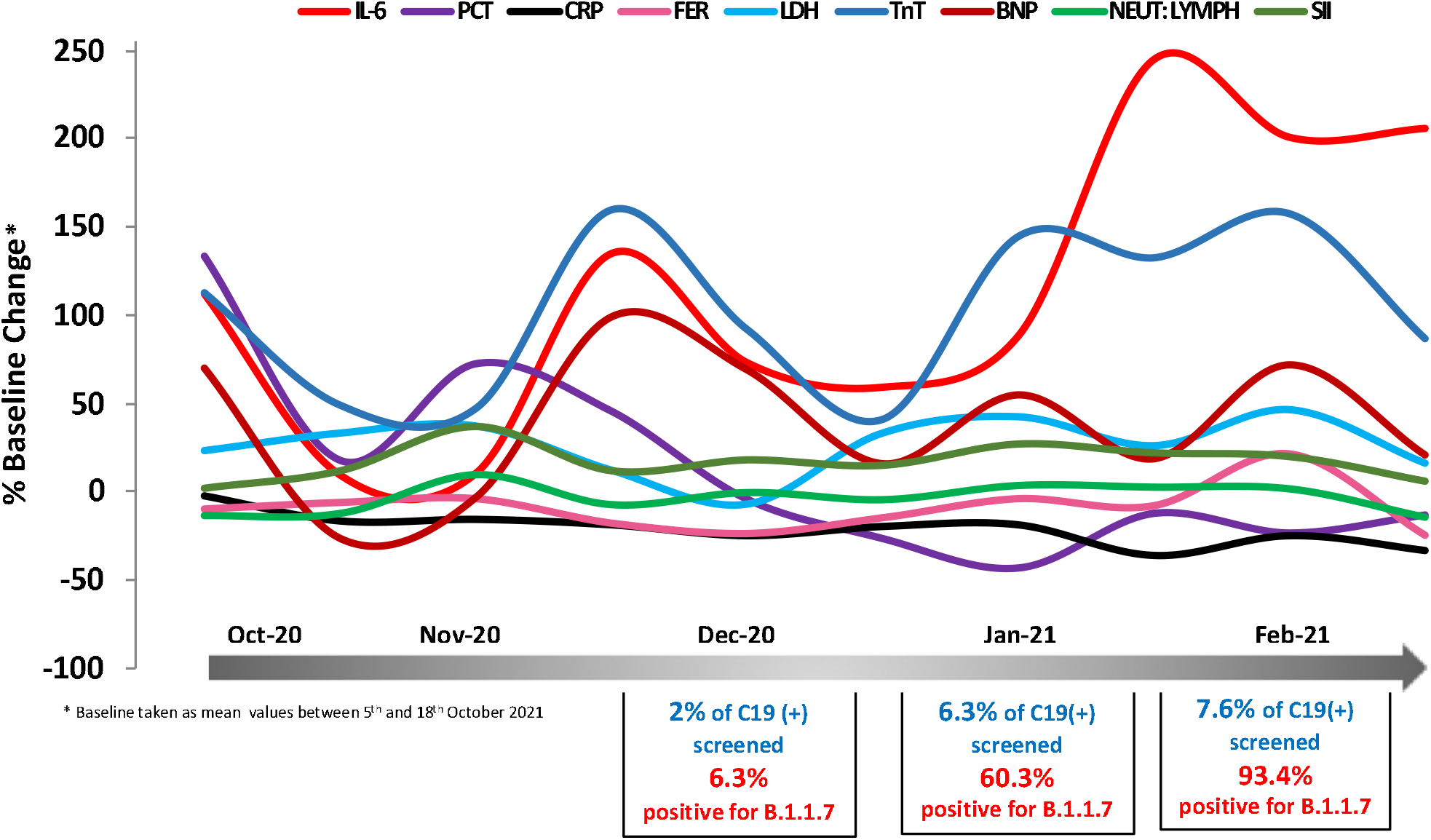
Temporal changes in blood COVID-19 biomarkers during the period 5/10/2020-7/3/2021. Mean values expressed as % change from baseline (mean value of 5^t^-18^th^ October 2020) are plotted as a function of time using measurements at regular time intervals (twice a month). Biomarkers of interest were measured at the routine Biochemistry laboratory of UHCW NHS Trust using CE-marked Elecsys kits from Roche Diagnostics. In total 1,112 data points were analysed.

### COVID-19 biomarker levels in deceased vs survivors as a function of *alpha*/B.1.1.7 presence

This data led us to explore the COVID-19 biomarker profile in hospitalised patients infected with or without the *alpha*/B.1.1.7 VOC. We focused our analysis on a small cohort of COVID-19 patient samples where *alpha*/B.1.1.7 VOC status was either confirmed by NGS or obtained from a time period without any *alpha*/B.1.1.7 VOC cases (25) (positive for B.1.1.7 n=44; negatives n=39), and COVID-19 biomarkers and clinical outcomes were available. The *alpha*/B.1.1.7 (+) group had a survivor: deceased ratio of 3:1, whereas in the *alpha*/B.1.1.7(-) group the ratio was 1:1.

Patients were categorised according to outcome (survived *vs deceased*) and *alpha*/B.1.1.7 presence or absence. Data analysis and biomarker comparison across groups was based on calculation of modified z-scores for each biomarker to identify variation from the group median: results showed that for both *alpha*/B.1.1.7 positive and negative groups, deceased patients had raised modified z-scores for several biomarkers investigated whereas survivors had generally negative z-scores indicative of lower values (Figure 4). As expected IL-6 exhibited the most marked difference in *alpha*/B.1.1.7 (+) deceased vs survivors followed by LDH. In both *alpha*/B.1.1.7 (+) and (-) groups, there was a clear difference in the modified z-scores of most inflammatory biomarkers including CRP, N/L ratio (as well as pro-NTBNP and cTnT) between deceased vs survivors, pointing towards a more potent inflammatory response in the deceased and activation of multiple biomarkers and pathogenic pathways. Comparing *alpha*/B.1.1.7(+) and (-), LDH and SII were greater than the overall group median in *alpha*/B.1.1.7(+) patients, even in those with favourable outcomes

**Fig.4.**
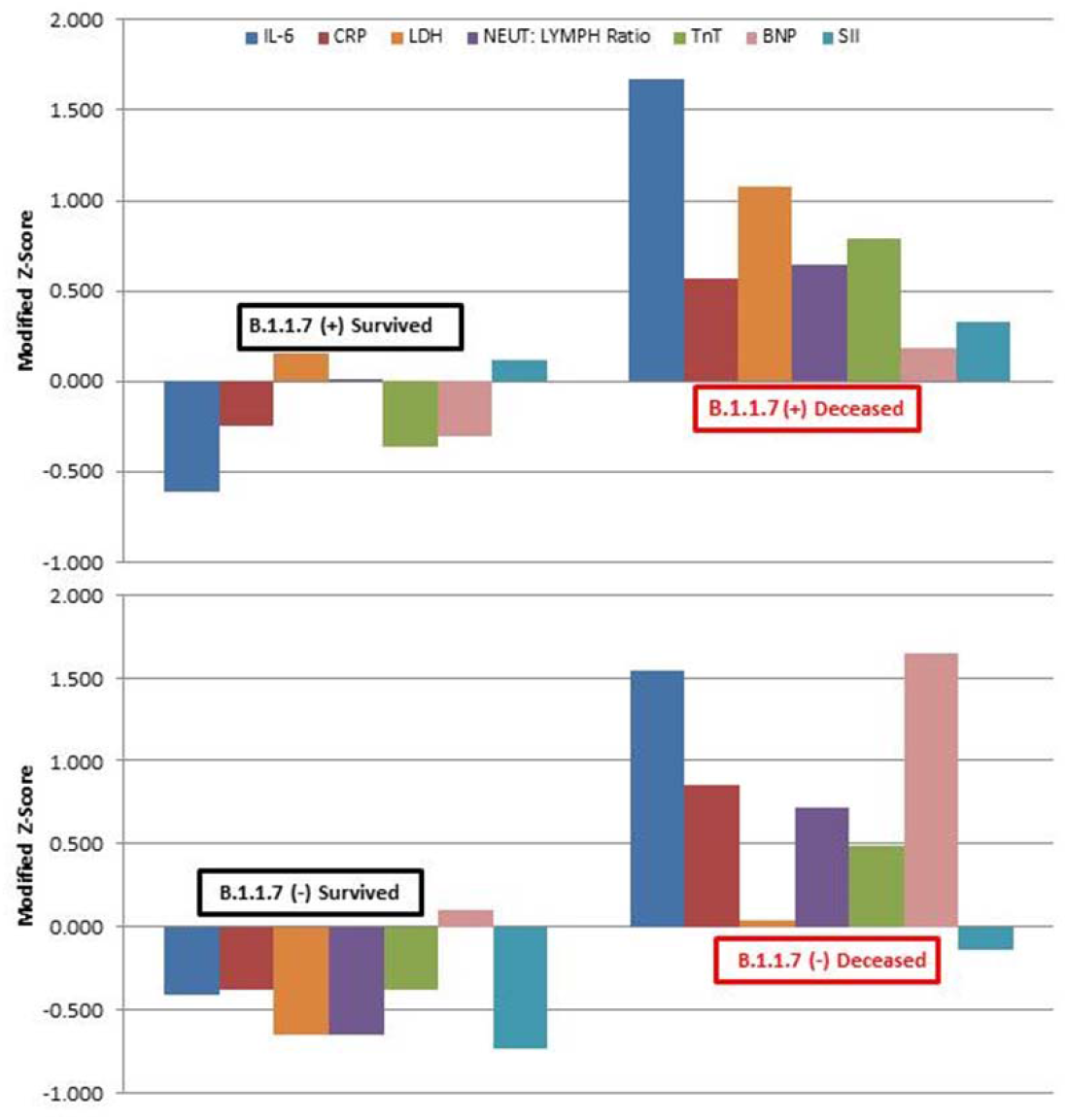
Selected blood COVID-19 biomarker levels in hospitalised patients with confirmed *alpha*/B.1.1.7 status. Data is presented as z-scores to demonstrate deviation from group mean values. In total 83 patients’ data were analysed (positive for *alpha*/B.1.1.7 n=44; negatives n=39).

## DISCUSSION

Following identification of the first case in South East of England in Sept 2020, the *alpha*/B.1.1.7 VOC spread rapidly across the UK and with more than 30,000 probable cases by February 2021, it became the dominant SARS-CoV-2 variant in the UK and elsewhere (26). Initial approaches to monitor its transmission rates were based on the finding that specific qPCR assays exhibited S gene target failure (SGTF) due to presence of Δ69-70 that prevented S gene qPCR probe annealing (27). This proxy marker for this variant was used primarily in Pillar 2 labs employing Δ69-70-sensitive PCR assays. However, this approach was not suitable for Pillar 1 NHS diagnostic labs interested in monitoring presence of *alpha*/B.1.1.7 VOC amongst hospitalised patients that employ a wide variety of PCR methods often insensitive to Δ69-70. To address this gap, we used a PCR protocol based on sequential detection of N501Y and Δ69-70 in Covid-19 positive samples. Although the N501Y is present in multiple variants including *alpha*/B.1.1.7 (UK or Kent), *beta*/B.1.351 (South Africa) and *gamma*/P1 (Brazil), the combination of N501Y and Δ69-70 appears to be *alpha*/B.1.1.7 -specific, at least at present until the emergence of other variants. Full concordance with sequencing data confirmed that this PCR protocol can provide an alternative rapid approach suitable for triaging samples that require additional confirmatory sequencing.

The mutation assays employed genotyping based on melting temperature shifts. Analytically, this protocol is suitable for routine use as it can be implemented in labs without expensive sequencing platforms and highly skilled staff. Moreover, these assays demonstrated appropriate performance characteristics, in terms of analytical sensitivity and specificity, RNA quality requirements, repeatability and reproducibility. In our hands the failure rate of the N501Y assay was not negligible, and this was observed only in specimens with low viral loads based on borderline Ct values of the original RealT*ime* SARS-CoV-2 assay. As this was a retrospective analysis, possible sample deterioration cannot be excluded. Nevertheless, this identifies a possible limitation of the assay that requires careful monitoring of specimen handling and storage.

Application of this screening protocol in a random representative pool of positive samples tested in an NHS Pillar 1 laboratory during the SARS-CoV-2 second wave of winter 2020-2021 suggested that the *alpha*/B.1.1.7 VOC became the dominant variant early in January 2021 in our Pillar 1 laboratory. Earlier reports raised the possibility of this variant being associated with increased risk of death compared with other variants (15-17) as some of the mutation may increase ACE-2 receptor binding affinity. Although monthly death rates in hospitalised patients could not provide clear evidence of consistently and sustained increased mortality due to *alpha*/B.1.1.7 VOC, we did monitor levels of various biomarkers associated with COVID-19 severe disease (28, 29).

During January-February of 2021, the period during which the prevalence of *alpha*/B.1.1.7 increased most notably, the mean levels of IL-6 were significantly higher compared to the previous 14-week mean. This unpredicted finding raises the possibility that hospitalised patients infected with the *alpha*/B.1.1.7 variant exhibited more severe disease. IL-6 has previously been proposed as a biomarker indicating excessive inflammation and disease severity (30, 31). Although the mortality rates during January-February of 2021 were not notably different, admitted patients during that period were generally younger (67 *vs* 63 years). However, firm conclusions about a direct causative link between *alpha*/B.1.1.7 VOC and disease severity require availability of detailed clinical data and potential confounders. To address this, we analysed blood biomarker data in a small group of patients with confirmed the *alpha*/B.1.1.7 VOC status. This is the first study that correlates directly biomarker levels to SARS-CoV-2 variants. A clear pattern emerged that deceased patients in both *alpha*/B.1.1.7 (+) or (-) groups had biomarkers above the group mean whereas survivors had lower than the mean levels. The two deceased groups had some notable differences and highest z-scores were seen in IL-6 and LDH levels in the *alpha*/B.1.1.7 (+) deceased group indicative of extensive disease burden characterised by hyperinflammation and increased tissue damage. This finding might be of prognostic value as IL-6 is an established therapeutic target of COVID-19 (32-34) and a recent study reported reduced effectiveness of IL-6 inhibition and increased mortality in patients with high LDH concentrations. In contrast, only moderate changes in these two biomarkers was observed in the *alpha*/B.1.1.7(-) deceased group, although this group had relative higher z-scores for CRP, an acute phase protein.

There is conflicting evidence whether *alpha*/B.1.1.7 VOC infections are associated with increased risk of death; although earlier studies (15-17) reported increased risk of death and estimated an increased hazard ratio for the risk of death of 61–67% for the *alpha*/B.1.1.7 VOC using individual-level data, a recent ecological study found no changes in reported symptoms or disease duration associated with *alpha*/B.1.1.7 (35). Similar findings have been reported in other studies that used early population-level data but were unable to demonstrate differences in mortality when individual-level data were used (25, 36).

Our inpatient data suggest possible manifestation of a more severe disease phenotype associated with IL-6 induced hyperinflammation. This finding is of clinical significance as IL-6 remains the best available biomarker for severity of COVID-19 and is considered by many, the most promising marker for guiding treatment. Larger multicentre studies and access to detailed clinical data including *alpha*/B.1.1.7 status is required to establish whether *alpha*/B.1.1.7 VOC can indeed drive more severe inflammatory phenotypes and ultimately lethal disease in addition to enhanced infectivity/transmissibility and ability to circumvent drug and immune control.

In summary, innovative use of mutation detection assays can provide a suitable tool for SARS-CoV-2 variants surveillance. In fact, this approach is been developed by various diagnostics services worldwide. Rapid deployment of such tools can support efforts to contain transmission of variants especially VOCs with increased rates of transmissibility that can lead to hospitalisation of COVID-19 patients with more severe disease phenotypes and possibly poor acute and long-term outcomes.

## Data Availability

Data used in this study can be made available upon request.

## ACKNOWLEDGMENTS

We would like to thank all UHCW Pathology staff and Garry Georgiades, Performance and Informatics Directorate, UHCW NHS Trust for generating, extracting and analysing data used in this work.

## Notes

**Funding** The work presented has been funded by the Institute of Precision Diagnostics and Translational Medicine, Pathology, UHCW NHS Trust, Coventry, UK.

**Competing Interest Statement** The authors have no conflict of interest or financial relationships relevant to the submitted work to disclose. No form of payment was given to anyone to produce the manuscript.

### Competing Interest Statement

The authors have declared no competing interest.

### Funding Statement

The work presented has been funded by the Institute of Precision Diagnostics and Translational Medicine, Pathology, UHCW NHS Trust, Coventry, UK.

### Author Declarations

UHCW NHS Trust COVID-19 Research Committee considered the research and approved ethics oversight exception

